# Macronutrient Intake and Carbohydrate Quality in Indian Adults With and Without Type 2 Diabetes: The Multicentre I-STARCH-1 Cross-Sectional Study

**DOI:** 10.1101/2025.11.04.25339356

**Authors:** Nitin Kapoor, Sanjay Kalra, Neeta Deshpande, Sheryl S. Salis, Smriti Gadia, Thamburaj Anthuvan

## Abstract

**Background/Objectives:** India’s diet remains dominated by carbohydrates; the recent ICMR– INDIAB survey-21 found that carbohydrates contribute approximately 62% of total energy intake nationally, alongside relatively low protein intake, but did not assess carbohydrate quality by diabetes status. I-STARCH-1 characterised macronutrient intake and carbohydrate quality among Indian adults with and without type 2 diabetes mellitus (T2DM), with descriptive comparison to the original STARCH (Study to Assess dietaRy CarboHydrate content) 2014 cohort.

**Methods:** This multicentre cross-sectional study enrolled 1104 adults (690 with T2DM and 414 without T2DM) from 29 healthcare centres across 14 Indian states. Dietary intake was assessed using three non-consecutive 24-hour recalls and harmonised using the Indian Council of Medical Research–National Institute of Nutrition (ICMR–NIN) Food Composition Tables. Macronutrient intake was compared by T2DM status and geographic zone, with STARCH 2014 used as a historical reference. Exploratory Diet-Quality Index (DQI) and Protein–Fat Quality Index (PFQI) scores were also evaluated.

**Results:** Carbohydrates contributed 62.1% of total energy intake, fat 25.1%, and protein 12.8%. Simple carbohydrates accounted for 20.3% of total carbohydrate intake, while fibre intake remained low (∼15 g/day). Adults with T2DM derived a lower proportion of energy from carbohydrates than those without T2DM (61.0% vs. 64.1%). Dietary patterns were broadly consistent across geographic zones. Compared descriptively with STARCH 2014, I-STARCH-1 showed lower carbohydrate intake, higher fat intake, a higher simple-carbohydrate proportion, and less favourable exploratory DQI and PFQI values. Convenience clinic-based sampling and the cross-sectional design limit generalisability and causal inference.

**Conclusions:** Carbohydrate intake among Indian adults remains high, while carbohydrate quality is suboptimal, characterised by low fibre and elevated simple-carbohydrate intake. Findings support prioritising carbohydrate quality, fibre, and protein adequacy rather than reducing carbohydrate quantity alone.

## 1. Introduction

India’s ongoing nutrition transition reflects a broader global shift in dietary patterns, characterized by changes in macronutrient balance and concerns regarding carbohydrate quality. Increasing evidence suggests that the metabolic effects of dietary carbohydrates depend not only on the quantity consumed but also on their quality, including the relative contributions of whole grains, dietary fiber, and simple sugars (Reynolds et al., 2019; Ma et al., 2024). Distinguishing carbohydrate quality from carbohydrate quantity has therefore become central to understanding diet-related metabolic risk. Type 2 diabetes mellitus (T2DM) remains a major global health challenge, projected to affect 783 million adults by 2045 (International Diabetes Federation, 2025).

Although dietary patterns vary considerably across India’s regions and cultures, many diets continue to derive a substantial proportion of total energy from cereal-based carbohydrates, particularly refined rice and wheat, while remaining relatively low in dietary fiber and protein density (Anjana et al., 2022; Green et al., 2016; ICMR–NIN, 2024; Joshi et al., 2014; Mohan et al., 2023a; Saboo et al., 2022). India bears a disproportionate burden of T2DM, with an estimated 101 million adults living with diabetes and 136 million with prediabetes in 2023 (Anjana et al., 2023). These dietary patterns are not uniform nationally; substantial regional, cultural, and socioeconomic variations exist in staple foods, meal composition, and food access across India, and this traditional pattern has been further altered by evolving food environments in recent years (Green et al., 2016; ICMR–NIN, 2024). Collectively, these features underscore the need for contemporary multicenter evidence on habitual macronutrient intake and carbohydrate quality in diverse Indian populations.

The STARCH (STudy to Assess dietaRy CarboHydrate content) study, conducted in 2014, was the first large multicenter dietary survey to characterize macronutrient intake among Indian adults with and without T2DM (Joshi et al., 2014). In one Indian analysis, carbohydrates contributed approximately 64% of total energy, exceeding the recommended 56%, and this was associated with less favorable metabolic outcomes. However, this value represents an observational association rather than a prescriptive dietary threshold and is consistent with the broader evidence linking higher carbohydrate intake to adverse cardiometabolic outcomes (Reynolds et al., 2019; Ma et al., 2024; Anjana et al., 2025). As the only comparable multicenter dataset for India, STARCH 2014 remains an important historical reference and informs the rationale for the present follow-up study (Kalra et al., 2025). However, dietary patterns and food environments have evolved substantially over the past decade, limiting the ability of a decade-old survey to accurately reflect contemporary dietary intake (Anjana et al., 2023; Paul & Paul, 2023).

Since the STARCH study (2014), India’s food and nutrition landscape has changed substantially. Rapid urbanisation, rising incomes, and the growth of digital food delivery have increased exposure to ready-to-eat, ultra-processed foods, packaged snacks, and sugar-sweetened beverages (Kumar et al., 2022; Malik & Hu, 2022; Murthy et al., 2025; Paul & Paul, 2023). Concurrent changes in the availability, accessibility, and affordability of pulses (John et al., 2021), updated national dietary guidance (ICMR–NIN, 2024; Hemalatha et al., 2023), and advances in diabetes management (Mohan et al., 2023b) may have further influenced habitual macronutrient intake. Emerging work characterising the carbohydrate profile and glycaemic index of traditional Indian foods (Shobana et al., 2022) has refined understanding of carbohydrate quality, reinforcing the need for an updated multicentre assessment capturing both the quantity and quality of contemporary dietary intake in India.

Composite diet-quality indices are widely used in nutritional epidemiology to summarize multiple dimensions of dietary intake into a single interpretable measure (Chiuve et al., 2012; Guenther et al., 2013). In the present study, the Diet Quality Index (DQI) and Protein–Fat Quality Index (PFQI) were investigator-derived composite measures developed to summarize macronutrient balance, including protein adequacy, fiber density, and simple carbohydrate exposure. These indices were used solely as descriptive summary measures. They are not established diagnostic or prognostic tools, have not undergone external validation, and should therefore be interpreted as exploratory measures rather than as validated dietary assessment instruments.

Despite the growing diabetes burden and rapidly changing food environment in India, contemporary multicenter evidence characterizing macronutrient distribution, carbohydrate quality, and regional variation among Indian adults with and without T2DM remains limited. I-STARCH-1 was conducted to address this gap. The primary objective was to characterize contemporary macronutrient intake and carbohydrate quality among Indian adults with and without T2DM across four geographic zones; secondary objectives were to compare patterns by diabetes status and zone, benchmark findings against STARCH 2014, and explore dietary balance using the investigator-derived DQI and PFQI. We hypothesized that the contemporary cohort would demonstrate poorer carbohydrate quality than the published STARCH 2014 cohort.

## 2. Methodology

### 2.1 Study design and setting

The Indian Study to Assess Real-World Carbohydrate Consumption (I-STARCH-1) was a multicenter, cross-sectional observational study conducted in 2025 across 29 participating healthcare centers located in 14 Indian states, representing the north, south, east, and west geographic zones of India. The study protocol has been published previously; detailed operational procedures, investigator training, and study workflow are described therein and summarized here for completeness (Kalra et al., 2025). Reporting followed the STROBE statement for observational studies, extended by the STROBE-nut recommendations for nutritional epidemiology (von Elm et al., 2007; Lachat et al., 2016); the completed STROBE-Nut checklist is provided in Supplementary Table S1. Participants were recruited through outpatient clinics and participating hospitals using convenience sampling; therefore, the sample was geographically diverse but not representative of the population. All centers implemented standardized protocols for recruitment, dietary assessment, investigator training, and data quality assurance under central coordination. The final analytic sample comprised 1,104 adults (690 with T2DM and 414 without). The study workflow, from setup through data harmonization, statistical analysis, and comparison with the STARCH 2014 dataset, is detailed in the Methods subsections that follow.

### 2.2. Ethical Approval

The study was approved by the Institutional Ethics Committee of Shah Lifeline Hospital & Heart Institute (Approval No. SLH-IEC/2025/006; approved on January 17, 2025). The study was conducted in accordance with the Declaration of Helsinki, the ICMR National Ethical Guidelines for Biomedical and Health Research Involving Human Participants, International Council for Harmonisation Good Clinical Practice (ICH-GCP) guidelines, and other applicable regulatory requirements. As a non-interventional, cross-sectional observational study, I-STARCH-1 did not require registration under the New Drugs and Clinical Trials (NDCT) Rules, 2019. Written informed consent was obtained from all participants prior to enrolment. The study protocol was published previously (Kalra et al., 2025).

### 2.3. Participants and Eligibility

Stratification by T2DM status was central to the study’s primary objective of characterising carbohydrate quality differences between adults with and without diabetes, given the direct relevance of macronutrient composition to glycaemic management in T2DM. Adults aged ≥18 years, with or without T2DM, were recruited from outpatient clinics, hospitals, and community programs across the 14 participating states. No upper age limit was specified; the observed age range of the enrolled participants was 19–88 years (mean age 49.8 ± 11.9 years). All participants provided written informed consent, and their medical records were available within the preceding 3 months. Eligibility for the T2DM and non-T2DM groups followed predefined clinical criteria, with additional exclusions based on medical conditions, lifestyle factors, and data quality. The detailed inclusion and exclusion criteria are listed in Table 1. Participants with incomplete or unreliable dietary recall data were excluded from the final analytical sample, which comprised 1,104 adults (690 with T2DM and 414 without T2DM).

**Table 1.** Eligibility criteria for study participants.

| Category | Criteria |
| --- | --- |
| Inclusion – General | Adults aged $\geq 18$ years; provided written informed consent; medical records available within the preceding 3 months; clinically stable for $\geq 30$ days. |
| Inclusion – T2DM | Confirmed diagnosis of T2DM according to American Diabetes Association (ADA) criteria; disease duration $\geq 6$–12 months; stable antidiabetic medication for $\geq 3$ months; no hospitalisation within the preceding 3 months. |
| Inclusion – Non-T2DM | No history of diabetes mellitus; fasting plasma glucose $< 126$ mg/dL or HbA1c $< 6.5\%$; not receiving glucose-lowering medication; not following medically prescribed restricted diets. |
| Exclusion – General | Pregnancy or lactation; severe cognitive impairment; active malignancy or terminal illness; hospitalisation within the preceding 30 days; incomplete or unreliable dietary data. |
| Exclusion – Medical | Type 1 diabetes or secondary diabetes; chronic kidney disease (CKD; eGFR $< 30$ mL/min/1.73 m<sup>2</sup>); active liver disease or cirrhosis; eating disorders. |
| Exclusion – Lifestyle | Participation in structured weight-loss programmes; major dietary change within the preceding 3 months; inability to complete dietary recalls. |

**Note:** Age eligibility was $\geq 18$ years, with no upper age limit specified. The observed age range of enrolled participants was 19–88 years (mean age $49.8 \pm 11.9$ years). **Abbreviations:** ADA, American Diabetes Association; T2DM, type 2 diabetes mellitus; HbA1c, glycated haemoglobin; CKD, chronic kidney disease; eGFR, estimated glomerular filtration rate.

### 2.4. Definition of Type 2 Diabetes Mellitus

T2DM status was determined using documented medical records or, where documentation was unavailable, self-reports supported by current glucose-lowering medication use. Where available, biochemical records (fasting plasma glucose and/or HbA1c) obtained within the preceding three months were used to support the classification. Participants in the T2DM group had a physician-confirmed diagnosis according to the American Diabetes Association (ADA) criteria and were receiving stable glucose-lowering therapy. Participants in the non-T2DM group had no documented history of diabetes, were not receiving glucose-lowering medication, and met the glycemic eligibility criteria (fasting plasma glucose <126 mg/dL or HbA1c <6.5%) where such data were available. Individuals with type 1 and secondary diabetes were excluded on clinical grounds.

### 2.5. Recruitment and Participant Flow

Participants were recruited from outpatient clinics and participating hospitals across 29 healthcare centers in 2025 using convenience sampling. The number of individuals approached before formal screening was not recorded consistently across centers; therefore, a conventional response rate (enrolled divided by approached) could not be calculated. Of the 1,358 individuals screened against the predefined eligibility criteria, 1,137 (83.7%) were eligible and enrolled, the closest available indicator of recruitment efficiency in the absence of pre-screening approach data, while 221 were excluded for ineligibility, declined participation, or incomplete baseline information. Enrolment was well distributed across centers (median 40 per center; mean 39.2; range 19 to 50), with 26 of 29 centers meeting the target of 40; one center enrolled above target (n=50), and two enrolled below target (n=19 and n=27), likely reflecting site-level recruitment constraints. Following the data quality review, a further 33 participants were excluded due to incomplete or unreliable dietary recalls, implausible energy intake (below 800 or above 4,000 kcal/day), or recent participation in specialized diet programs (disaggregated counts not recorded). The final analytic sample comprised 1,104 adults (690 with T2DM and 414 without). The recruitment and selection processes are summarized in Figure 1.

**Figure 1.**
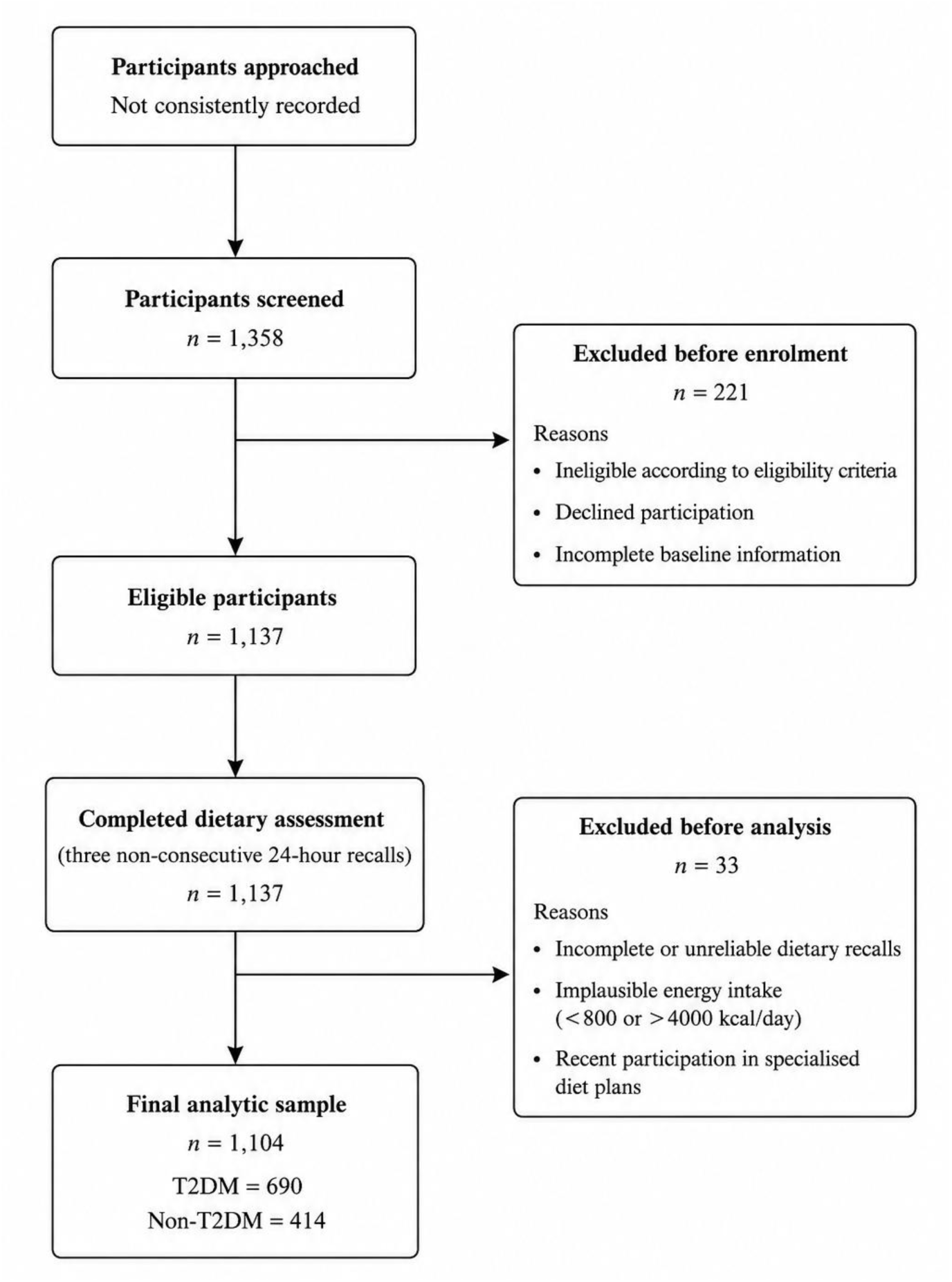
Participant recruitment and selection flow diagram for the I-STARCH-1 study.

### 2.6. Sample Size Determination

A formal a priori sample size or power calculation was not conducted. Instead, the study adopted a pragmatic recruitment strategy based on the protocol target of approximately 40 participants per center (approximately 30 with T2DM and 10 without), designed to achieve broad geographic coverage and balanced recruitment across diabetes strata. Of the 1,137 eligible participants, 1,104 were retained in the final analytic dataset following data quality exclusions. This sample provided high precision for the primary outcome: mean carbohydrate contribution (62.1% of total energy, standard deviation [SD] 5.4) was estimated with a 95% confidence interval of approximately ±0.3 percentage points, indicating adequate precision for the study’s descriptive and comparative objectives despite the absence of a formal power calculation.

### 2.7. Dietary Assessment

Habitual dietary intake was assessed using three non-consecutive 24-hour dietary recalls (two weekdays and one weekend day), administered face-to-face by trained investigators using a standardized multi-pass technique and calibrated household measures (Kalra et al., 2025). The recalls were conducted using a structured dietary survey questionnaire developed for the study, provided in Supplementary File S1. Field supervisors reviewed each recall for completeness and conducted random verification calls to address any discrepancies. The dietary intake from the three recalls was used to estimate the habitual intake for each participant. The interval between recalls and whether all recalls were completed within a predefined time window were not systematically recorded and, therefore, cannot be reported.

### 2.8. Food Composition and Nutrient Coding

All reported foods and beverages were coded using the ICMR–NIN Food Composition Tables. Each reported food item was matched to the most appropriate food entry for nutrient estimation. Mixed dishes were decomposed into their constituent ingredients based on the participant-reported recipe and preparation method, and the nutrient values were calculated by summing the corresponding ingredient-level values. Region-specific foods without a direct match were mapped using ingredient-level reconstruction or the nearest nutritionally comparable recipes. When manufacturer-specific nutrient information was unavailable for commercially packaged foods, branded products were matched to the closest generic food entry in the same food composition table. Energy and nutrient intakes were calculated for each recall day and averaged across the three dietary recalls to estimate the habitual intake.

### 2.9. Macronutrient Computation

Daily energy and nutrient intakes were calculated from coded dietary data using the ICMR–NIN Food Composition Tables. The total energy intake (kcal/day) was derived using standard Atwater conversion factors (4 kcal/g for carbohydrates, 4 kcal/g for protein, and 9 kcal/g for fat). Macronutrient intake was expressed as both absolute intake (g/day) and percentage contribution to total energy intake (%E). Dietary fiber was reported as g/day and g/1,000 kcal. Simple and complex carbohydrates were expressed as percentages of total carbohydrate intake, with complex carbohydrates calculated as the remaining carbohydrates after subtracting simple carbohydrates.

### 2.10. Study Outcomes

The primary outcomes were carbohydrate, protein, and fat intake as the percentage of total energy intake (%E), simple carbohydrate intake as the percentage of total carbohydrate intake, and dietary fiber intake (g/day and g/1,000 kcal). Secondary outcomes included comparisons between participants with and without T2DM, comparisons across the four geographic regions, and a descriptive comparison with the STARCH 2014 study. The investigator-derived Diet Quality Index (DQI) and Protein–Fat Quality Index (PFQI) were evaluated as exploratory outcomes because they have not undergone external validation.

### 2.11. Diet-Quality Index (DQI) and Protein–Fat Quality Index (PFQI)

The Diet-Quality Index (DQI) was calculated as:

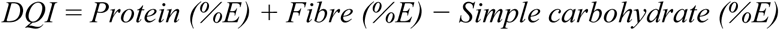

where all three components were expressed as the percentage of total energy intake (%E). Higher DQI values indicate better overall dietary quality. For this composite index only, fiber and simple carbohydrates were expressed as %E to maintain consistent units across all additive components. The DQI is an investigator-derived exploratory index informed by previous diet quality frameworks but has not undergone external validation and should not be interpreted as a validated diagnostic or prognostic measure (Chiuve et al., 2012; Guenther et al., 2013).

The Protein–Fat Quality Index (PFQI) was calculated as:

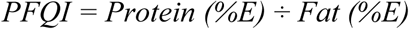

Higher PFQI values indicate a greater relative contribution of protein to fat intake. As no established clinical thresholds exist, the PFQI was used as an exploratory descriptive index, and its values should not be interpreted as indicating clinical benefit or harm.

### 2.12. Data Quality Assurance

All dietary records underwent standardized quality checks prior to analysis. Records were reviewed for completeness, internal consistency, and plausibility of reported dietary intakes. Incomplete dietary recalls, logically inconsistent records, and implausible total energy intakes based on predefined absolute plausibility thresholds were excluded from the final dataset. Primary dietary outcomes were completed for all participants in the final analytical sample. Biochemical covariates (fasting plasma glucose, post-prandial glucose, and HbA1c) had substantial missingness (42.7%, 46.1%, and 43.1%, respectively), reflecting incomplete availability of recent medical records. No imputation of missing data was performed, and all analyses were conducted using complete case data.

### 2.13. Statistical Analysis

Continuous variables are presented as mean ± standard deviation (SD) for normally distributed data or median (interquartile range [IQR]) for non-normally distributed data. Categorical variables are presented as frequencies and percentages. Distributional assumptions were assessed before selecting parametric or non-parametric tests. Comparisons between participants with and without T2DM and across geographic regions used the independent-samples *t*-test or one-way ANOVA for normally distributed variables, the Mann–Whitney *U* or Kruskal–Wallis test for non-normally distributed variables, and the chi-square test for categorical variables. Ninety-five percent confidence intervals (95% CIs) are reported for the primary dietary outcomes. Analyses were performed using IBM SPSS Statistics for Windows, Version 29.0 (IBM Corp., Armonk, NY, USA), with a two-sided *P* < 0.05 considered significant. Published STARCH 2014 summary statistics served as historical reference values; these comparisons are contextual, as the studies differ in sampling, participant composition and dietary assessment, and harmonised individual-level historical data were unavailable.

#### Deviations from the published protocol

recruitment was completed at 29 of the planned 33 centres; the planned multivariable regression, imputation and sensitivity analyses were not performed, so all comparisons are unadjusted complete-case analyses; subgroup analysis by urban versus semi-urban setting was replaced by comparison across four geographic zones; and biochemical markers are not reported as outcomes owing to substantial missingness

## 3. Results

### 3.1. Baseline Characteristics

The baseline demographic and lifestyle characteristics of the 1,104 participants are summarized in Table 2. The study included 690 (62.5%) participants with T2DM and 414 (37.5%) participants without T2DM. The overall mean age was 49.8 ± 11.9 years (range, 19–88 years), and the cohort comprised 541 (49.0%) men and 563 (51.0%) women. The overall mean body mass index (BMI) was 24.3 ± 3.6 kg/m², and the mean estimated waist circumference was 92.2 ± 10.6 cm. Compared with participants without T2DM, those with T2DM were significantly older (51.3 ± 11.2 vs. 47.3 ± 12.6 years; P < 0.001), had significantly greater estimated waist circumference (93.0 ± 10.3 vs. 90.8 ± 10.8 cm; P < 0.001), and were more likely to report following a restricted diet during the preceding year (55.7% vs. 43.0%; P < 0.001). BMI did not differ significantly between the groups (24.4 ± 3.6 vs. 24.1 ± 3.6 kg/m²; P = 0.183). The geographic distribution differed marginally between the groups (P = 0.055). No statistically significant between-group differences were observed in sex, frequency of meals eaten outside the home or obtained through home delivery, daily meal frequency, daily water intake, exercise participation, or alcohol consumption. T2DM duration was not included in the study dataset.

**Table 2.** Baseline Demographic and Lifestyle Characteristics of the Study Participants (N = 1,104)

| Characteristic | Overall (N = 1,104) | T2DM (n = 690) | Non-T2DM (n = 414) | P value |
| --- | --- | --- | --- | --- |
| Age (years), mean ± SD | 49.8 ± 11.9 | 51.3 ± 11.2 | 47.3 ± 12.6 | <0.001 |
| BMI (kg/m<sup>2</sup>), mean ± SD | 24.3 ± 3.6 | 24.4 ± 3.6 | 24.1 ± 3.6 | 0.183 |
| Estimated waist circumference (cm), mean ± SD | 92.2 ± 10.6 | 93.0 ± 10.3 | 90.8 ± 10.8 | <0.001 |
| **Gender, n (%)** |  |  |  | **0.504** |
| • Male | 541 (49.0) | 344 (49.9) | 197 (47.6) |  |
| • Female | 563 (51.0) | 346 (50.1) | 217 (52.4) |  |
| Outside meals (times/week), mean ± SD | 1.6 ± 1.3 | 1.6 ± 1.3 | 1.7 ± 1.4 | 0.253 |
| Daily meal frequency, mean ± SD | 3.5 ± 1.0 | 3.5 ± 0.9 | 3.6 ± 1.0 | 0.169 |
| Daily water intake (L/day), mean ± SD | 2.1 ± 0.8 | 2.1 ± 0.8 | 2.2 ± 0.9 | 0.320 |
| **Geographic region, n (%)** |  |  |  | **0.055** |
| • North | 220 (19.9) | 140 (20.3) | 80 (19.3) |  |
| • South | 328 (29.7) | 223 (32.3) | 105 (25.4) |  |
| • East | 343 (31.1) | 203 (29.4) | 140 (33.8) |  |
| • West | 213 (19.3) | 124 (18.0) | 89 (21.5) |  |
| Restricted diet during the past year, n (%) | 562 (50.9) | 384 (55.7) | 178 (43.0) | <0.001 |
| Exercise participation, *n* (%) | 497 (45.0) | 318 (46.1) | 179 (43.2) | 0.293 |
| Alcohol consumption, *n* (%) | 184 (16.7) | 113 (16.4) | 71 (17.1) | 0.802 |

*Note: Data are presented as mean ± SD or *n* (%). *P* values were derived using independent-samples *t*-tests for continuous variables and chi-square tests for categorical variables. Restricted diet was defined as participant-reported deliberate limitation of specific foods, nutrients, or total dietary intake during the preceding year, whether self-directed or advised by a healthcare professional. Waist circumference values were estimated. T2DM duration was not captured. **Abbreviations:** BMI, body mass index; SD, standard deviation; T2DM, type 2 diabetes mellitus.*

### 3.2. Overall Energy and Macronutrient Intake

The mean daily energy intake was 1,788 ± 565 kcal/day; the possibility of recall-related under-reporting is considered a limitation. Carbohydrates contributed 62.1% of the total energy intake, followed by fat (25.1%) and protein (12.8%) (Figure 2A). The mean dietary fiber intake was 15.4 ± 3.2 g/day (9.1 ± 3.2 g/1,000 kcal). Simple carbohydrates accounted for 20.3% of the total carbohydrate intake, whereas complex carbohydrates accounted for 79.7% (Figure 2B).

**Figure 2.**
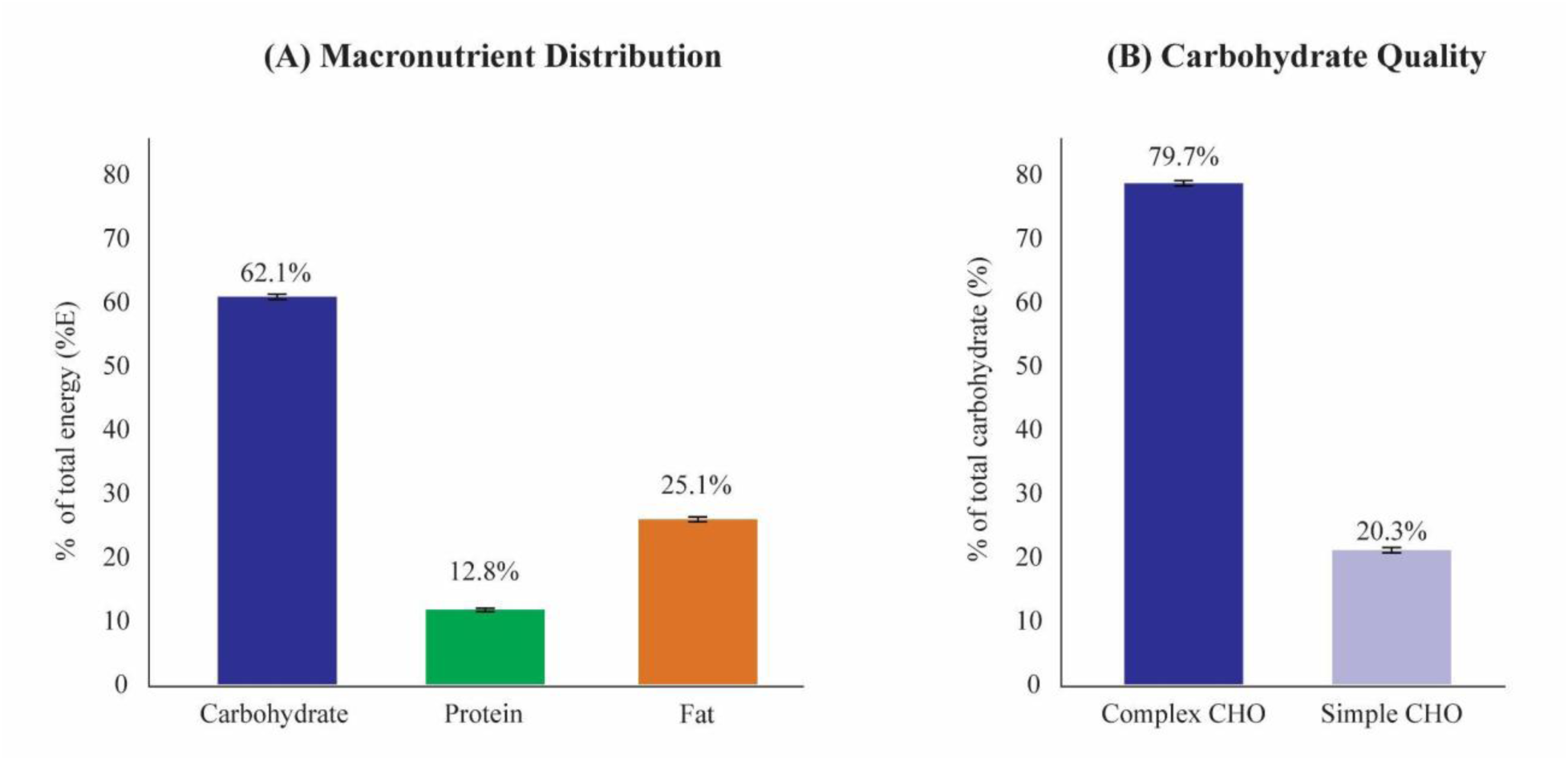
Overall Energy and Macronutrient Distribution in the I-STARCH 2025 Cohort **(A)** Mean percentage contribution of carbohydrate, protein, and fat to total energy intake among all study participants (N = 1,104). **(B)** Mean proportions of simple and complex carbohydrates, expressed as percentages of total carbohydrate intake, among all study participants. Error bars represent 95% confidence intervals. ***Note:*** *Values represent cohort means derived from three non-consecutive 24-hour dietary recalls. Macronutrient contribution is expressed as the percentage of total energy intake (%E). Simple and complex carbohydrates are expressed as percentages of total carbohydrate intake. **Abbreviations:** CHO, carbohydrate; %E, percentage of total energy intake; CI, confidence interval*.

### 3.3. T2DM Versus Non-T2DM

Participants with T2DM derived a lower proportion of total energy from carbohydrates and higher proportions from protein and fat than participants without T2DM (all P < 0.001; Table 3). Carbohydrate quality also differed between groups, with participants with T2DM consuming a lower proportion of simple carbohydrates and a correspondingly higher proportion of complex carbohydrates (both P < 0.001). The mean daily energy intake was lower among participants with T2DM, whereas dietary fiber intake did not differ significantly between the groups (P = 0.07).

**Table 3.**
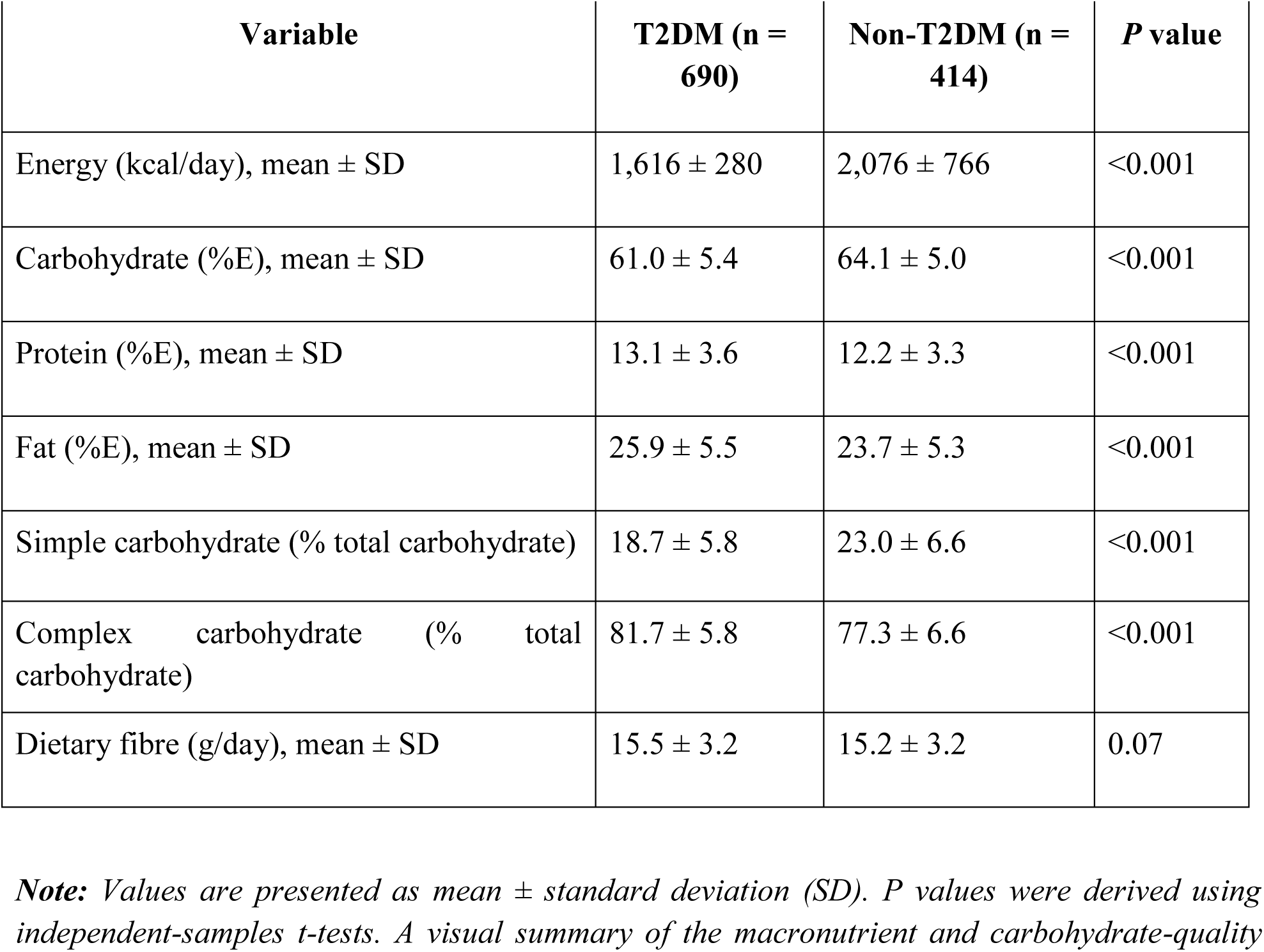
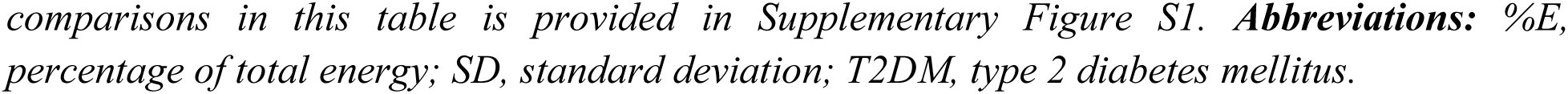
Macronutrient Profile by Diabetes Status in the I-STARCH 2025 Cohort.

| Variable | T2DM (n = 690) | Non-T2DM (n = 414) | P value |
| --- | --- | --- | --- |
| Energy (kcal/day), mean ± SD | 1,616 ± 280 | 2,076 ± 766 | <0.001 |
| Carbohydrate (%E), mean ± SD | 61.0 ± 5.4 | 64.1 ± 5.0 | <0.001 |
| Protein (%E), mean ± SD | 13.1 ± 3.6 | 12.2 ± 3.3 | <0.001 |
| Fat (%E), mean ± SD | 25.9 ± 5.5 | 23.7 ± 5.3 | <0.001 |
| Simple carbohydrate (% total carbohydrate) | 18.7 ± 5.8 | 23.0 ± 6.6 | <0.001 |
| Complex carbohydrate (% total carbohydrate) | 81.7 ± 5.8 | 77.3 ± 6.6 | <0.001 |
| Dietary fibre (g/day), mean ± SD | 15.5 ± 3.2 | 15.2 ± 3.2 | 0.07 |

**Note:** Values are presented as mean ± standard deviation (SD). P values were derived using independent-samples t-tests. A visual summary of the macronutrient and carbohydrate-quality

### 3.4. Geographic Distribution of Macronutrient Intake

The macronutrient distribution differed modestly across India’s four geographic zones (North, n = 220; South, n = 328; East, n = 343; West, n = 213) (Table 4). Carbohydrates remained the predominant source of dietary energy in all regions, contributing 61.5–63.2% of the total energy intake, followed by fat (22.9–26.6%) and protein (11.9–13.9%). Although the overall differences in carbohydrate contribution and simple carbohydrate intake across geographic zones were statistically significant (both one-way ANOVA, P < 0.001), the absolute between-region differences were modest in this study. Carbohydrate quality also showed limited regional variation, with simple carbohydrates contributing 19.6–21.5% and complex carbohydrates 78.9– 80.9% of the total carbohydrate intake. Overall, the dietary macronutrient composition was consistent across geographic regions. A geographical visualization of regional macronutrient distribution is provided in Supplementary Figure S2, following the presentation style of the original STARCH study.

**Table 4.**
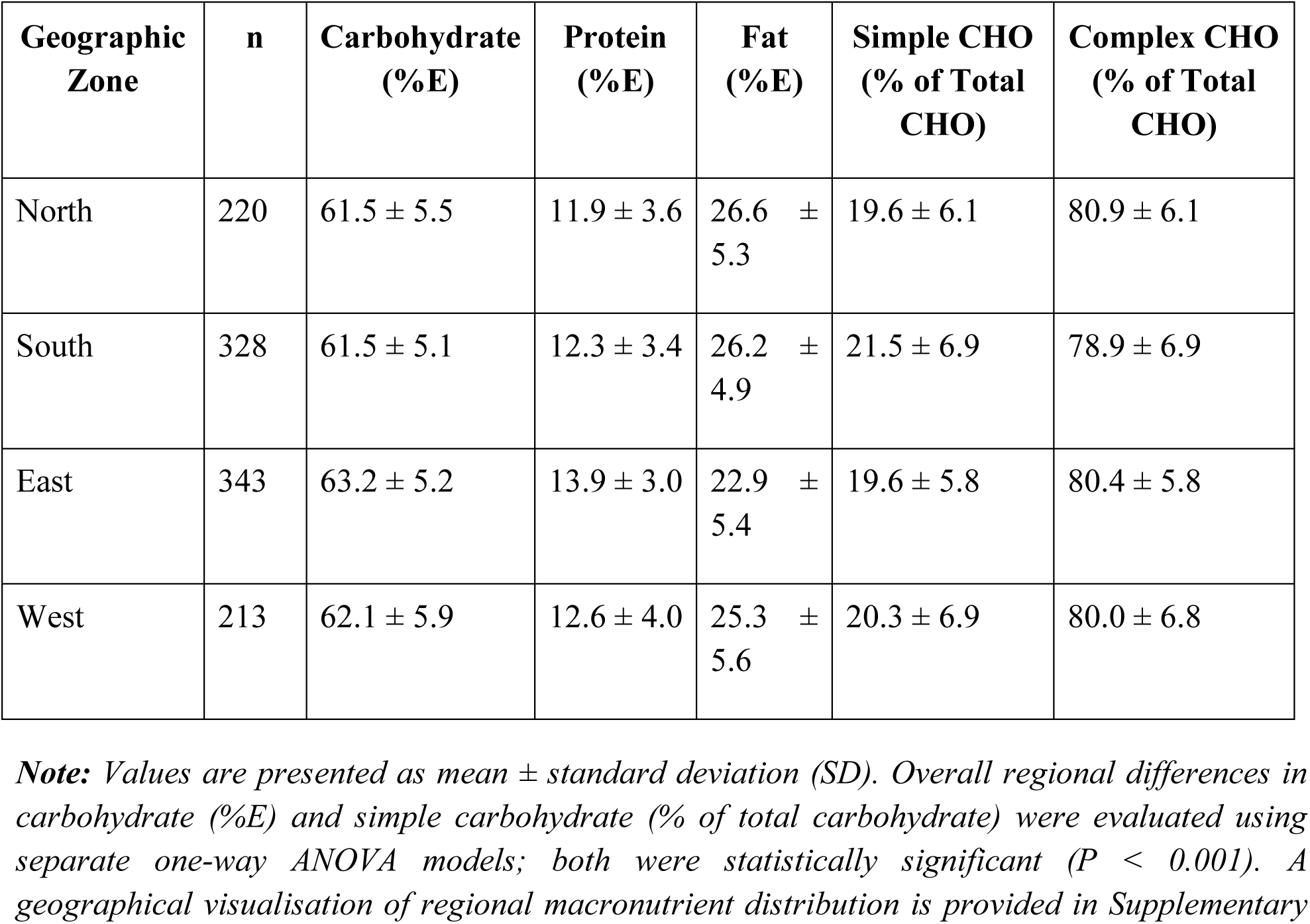
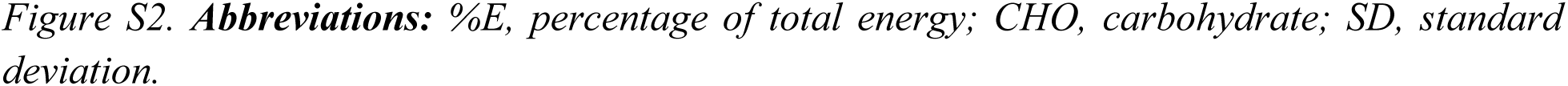
Regional Macronutrient Distribution in the I-STARCH 2025 Cohort.

| Geographic Zone | n | Carbohydrate (%E) | Protein (%E) | Fat (%E) | Simple CHO (% of Total CHO) | Complex CHO (% of Total CHO) |
| --- | --- | --- | --- | --- | --- | --- |
| North | 220 | 61.5 ± 5.5 | 11.9 ± 3.6 | 26.6 ± 5.3 | 19.6 ± 6.1 | 80.9 ± 6.1 |
| South | 328 | 61.5 ± 5.1 | 12.3 ± 3.4 | 26.2 ± 4.9 | 21.5 ± 6.9 | 78.9 ± 6.9 |
| East | 343 | 63.2 ± 5.2 | 13.9 ± 3.0 | 22.9 ± 5.4 | 19.6 ± 5.8 | 80.4 ± 5.8 |
| West | 213 | 62.1 ± 5.9 | 12.6 ± 4.0 | 25.3 ± 5.6 | 20.3 ± 6.9 | 80.0 ± 6.8 |

**Note:** Values are presented as mean ± standard deviation (SD). Overall regional differences in carbohydrate (%E) and simple carbohydrate (% of total carbohydrate) were evaluated using separate one-way ANOVA models; both were statistically significant ( $P < 0.001$ ). A geographical visualisation of regional macronutrient distribution is provided in Supplementary

### 3.5. Historical Comparison with the STARCH 2014 Cohort

Compared with the published STARCH 2014 cohort (Joshi et al., 2014), the I-STARCH 2025 cohort demonstrated a lower mean contribution of carbohydrates to total energy intake and a correspondingly higher contribution from fat, whereas protein intake differed only modestly. Among participants with T2DM, carbohydrate quality also differed, with a higher proportion of simple carbohydrates and a lower proportion of complex carbohydrates than those reported in STARCH 2014 (Table 5). Comparable historical carbohydrate-quality data for the non-T2DM and overall cohorts were unavailable because these metrics were not reported in the original STARCH study. As STARCH 2014 and I-STARCH 2025 represent independent cross-sectional cohorts conducted at different time points, these comparisons should be interpreted as descriptive rather than as evidence of temporal change.

**Table 5.**
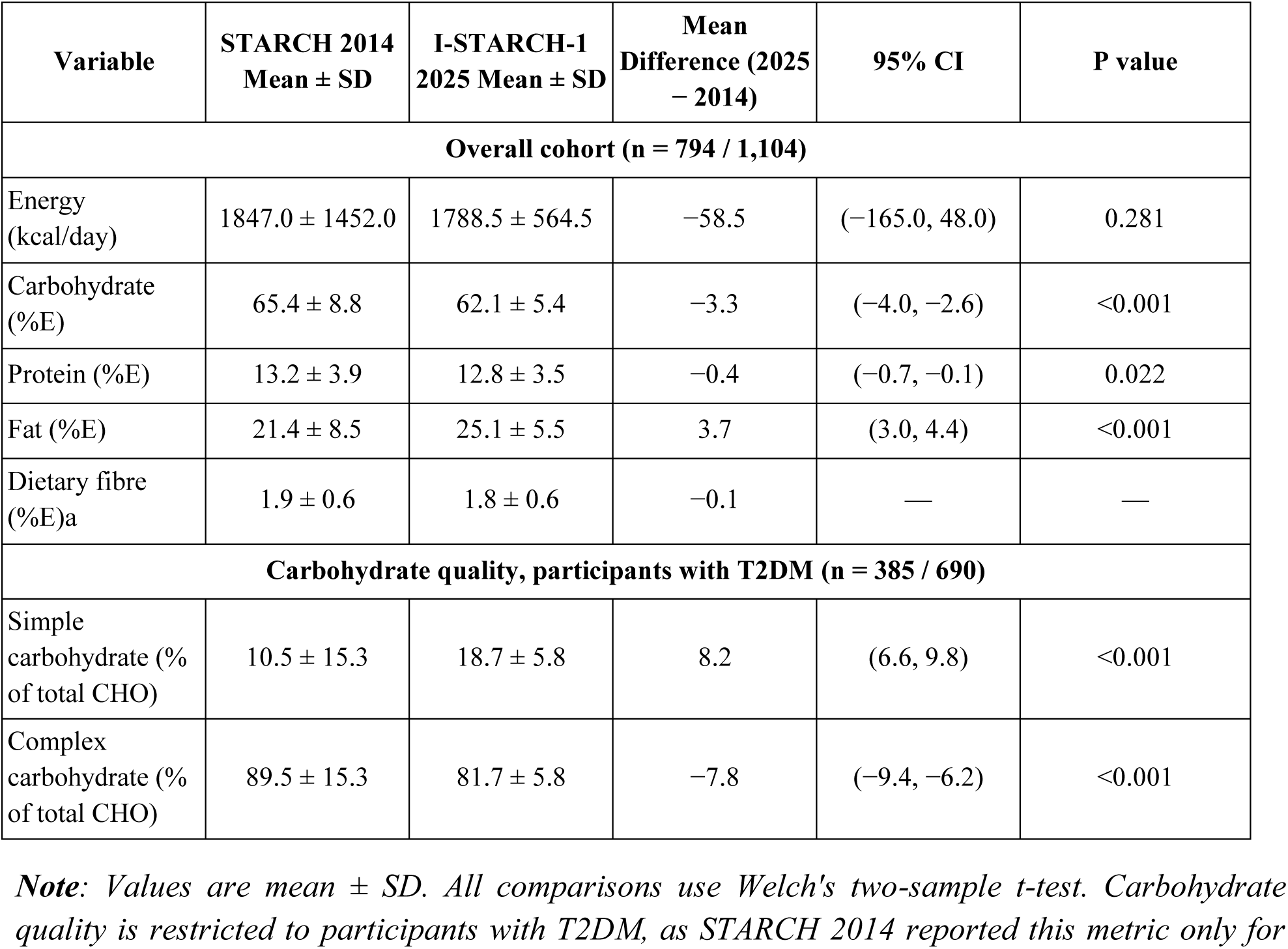
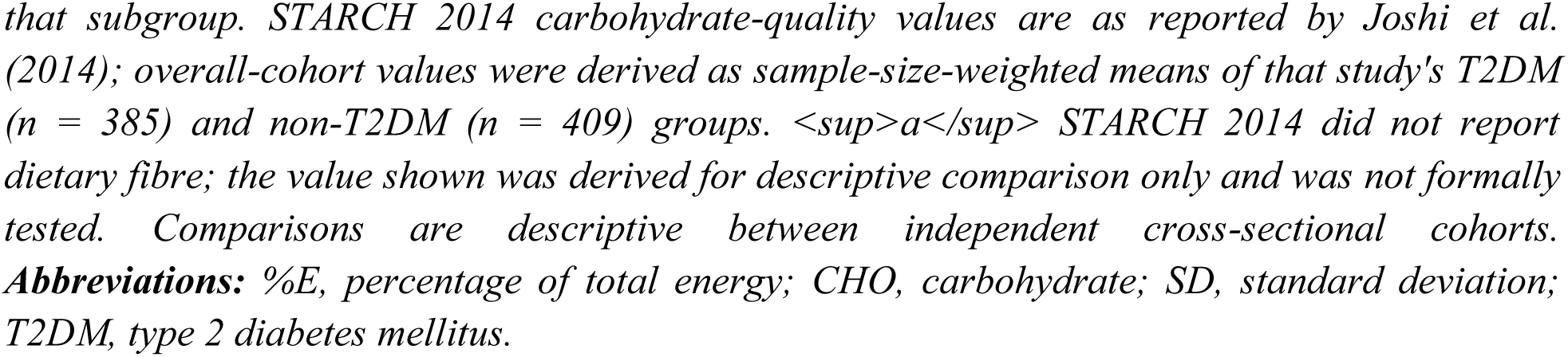
Descriptive Comparison Between the STARCH 2014 and I-STARCH-1 2025 Cohorts.

| Variable | STARACH 2014<br>Mean ± SD | I-STARACH-1<br>2025 Mean ± SD | Mean<br>Difference (2025<br>– 2014) | 95% CI | P value |
| --- | --- | --- | --- | --- | --- |
| **Overall cohort (n = 794 / 1,104)** |  |  |  |  |  |
| Energy (kcal/day) | 1847.0 ± 1452.0 | 1788.5 ± 564.5 | –58.5 | (–165.0, 48.0) | 0.281 |
| Carbohydrate (%E) | 65.4 ± 8.8 | 62.1 ± 5.4 | –3.3 | (–4.0, –2.6) | <0.001 |
| Protein (%E) | 13.2 ± 3.9 | 12.8 ± 3.5 | –0.4 | (–0.7, –0.1) | 0.022 |
| Fat (%E) | 21.4 ± 8.5 | 25.1 ± 5.5 | 3.7 | (3.0, 4.4) | <0.001 |
| Dietary fibre (%E)<sup>a</sup> | 1.9 ± 0.6 | 1.8 ± 0.6 | –0.1 | — | — |
| **Carbohydrate quality, participants with T2DM (n = 385 / 690)** |  |  |  |  |  |
| Simple carbohydrate (% of total CHO) | 10.5 ± 15.3 | 18.7 ± 5.8 | 8.2 | (6.6, 9.8) | <0.001 |
| Complex carbohydrate (% of total CHO) | 89.5 ± 15.3 | 81.7 ± 5.8 | –7.8 | (–9.4, –6.2) | <0.001 |

**Note:** Values are mean ± SD. All comparisons use Welch's two-sample t-test. Carbohydrate quality is restricted to participants with T2DM, as STARCH 2014 reported this metric only for
that subgroup. STARCH 2014 carbohydrate-quality values are as reported by Joshi et al. (2014); overall-cohort values were derived as sample-size-weighted means of that study's T2DM ( $n = 385$ ) and non-T2DM ( $n = 409$ ) groups. <sup>a</sup> STARCH 2014 did not report dietary fibre; the value shown was derived for descriptive comparison only and was not formally tested. Comparisons are descriptive between independent cross-sectional cohorts. **Abbreviations:** %E, percentage of total energy; CHO, carbohydrate; SD, standard deviation; T2DM, type 2 diabetes mellitus.

The historical differences in macronutrient distribution and carbohydrate quality are illustrated in Figure 3.

**Figure 3.**
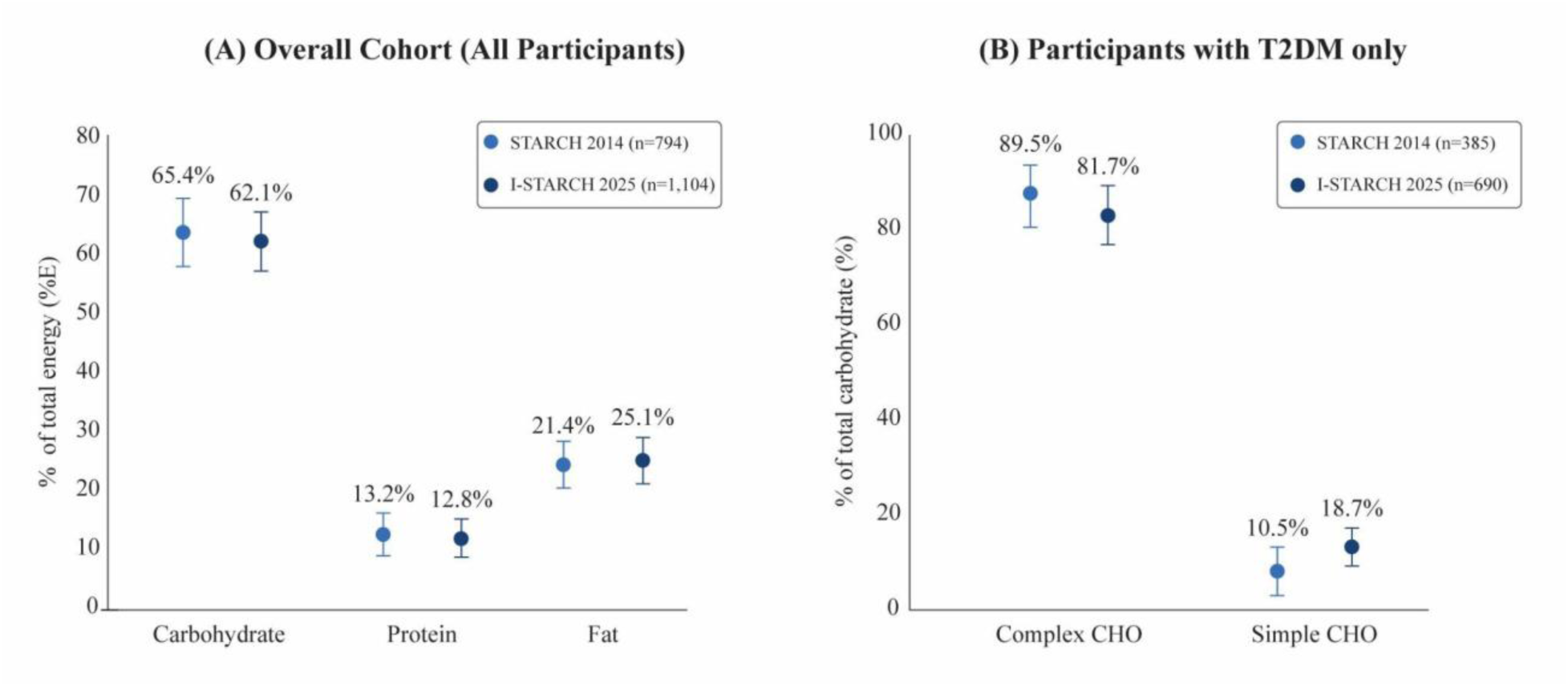
Historical Comparison of Macronutrient Distribution and Carbohydrate Quality Between STARCH 2014 and I-STARCH 2025 Cohorts A) Mean carbohydrate, protein, and fat contributions to total energy intake (%E) in the overall STARCH 2014 and I-STARCH 2025 cohorts. (B) Mean simple and complex carbohydrate contributions, expressed as percentages of total carbohydrate intake, among participants with T2DM in the STARCH 2014 and I-STARCH 2025 cohorts. Points represent cohort means and error bars represent 95% confidence intervals. ***Note:*** *STARCH 2014 and I-STARCH 2025 were independent cross-sectional cohorts and were not population matched. Panel A presents overall cohort comparisons, whereas Panel B presents T2DM-specific carbohydrate-quality comparisons because corresponding overall carbohydrate-quality measures were not reported in the original STARCH 2014 publication. Comparisons are descriptive and should not be interpreted as evidence of longitudinal or temporal change. **Abbreviations:** %E, percentage of total energy intake; CHO, carbohydrate; CI, confidence interval; T2DM, type 2 diabetes mellitus*.

### 3.6. Dietary Quality Indicators

Compared with the published STARCH 2014 cohort (Joshi et al., 2014), the I-STARCH 2025 cohort demonstrated a lower composite Diet Quality Index (DQI; protein %E + fibre %E − simple carbohydrate %E) (Table 6). This difference was driven primarily by a greater contribution of simple carbohydrates, while protein and dietary fibre (%E) were broadly similar between cohorts. Although dietary fibre contribution differed statistically (1.9%E vs. 1.8%E), the absolute difference was small and its nutritional significance is uncertain. As an investigator-derived, non-validated exploratory measure, the DQI should be viewed as indicating differences in overall macronutrient quality between cohorts rather than as confirmatory evidence of dietary deterioration. Given that the two studies represent independent cross-sectional cohorts at different time points, findings are descriptive and not evidence of temporal change.

**Table 6.** Comparison of Composite Diet-Quality Index Between STARCH 2014 and I-STARCH 2025.

| Variable | STARCH<br>2014 Mean ±<br>SD | I-STARCH<br>2025 Mean ±<br>SD | Mean<br>Difference<br>(2025 – 2014) | 95% CI | P value |
| --- | --- | --- | --- | --- | --- |
| Composite Diet-Quality Index<br>(DQI) | 7.7 ± 5.3 | 2.1 ± 6.2 | –5.6 | (–6.0,<br>–5.3) | <0.001 |

**Note:** Values are mean ± SD. DQI = protein (%E) + fibre (%E) – simple carbohydrate (%E); higher = better. STARCH 2014 components were derived from values reported by Joshi et al. (2014); the fibre component was derived rather than directly measured, as STARCH 2014 did not report dietary fibre, and should be considered exploratory. Comparisons are descriptive between independent cross-sectional cohorts. **Abbreviations:** DQI, Diet-Quality Index; %E, percentage of total energy; SD, standard deviation.

### 3.7. Protein–Fat Quality Index

Compared descriptively with the published STARCH 2014 cohort (Joshi et al., 2014), the Protein–Fat Quality Index (PFQI) was lower in the I-STARCH 2025 cohort (Table 7), indicating a lower protein-to-fat ratio. Similar descriptive differences were observed overall and among participants with and without T2DM. Because STARCH 2014 did not report PFQI stratified by diabetes status, both subgroup comparisons were referenced to the same overall STARCH 2014 PFQI value rather than matched subgroup-specific baseline values. Although all comparisons were statistically significant, no established clinical threshold exists for this investigator-derived index; results should therefore be interpreted descriptively rather than as evidence of clinically meaningful differences. As the two studies represent independent cross-sectional cohorts at different time points, findings should not be taken as evidence of temporal change.

**Table 7.** Comparison of Protein–Fat Quality Index Between STARCH 2014 and I-STARCH 2025.

| Group | STARCH 2014<br>Mean ± SD | I-STARCH<br>2025 Mean ± SD | Mean Difference<br>(2025 – 2014) | 95% CI | P value |
| --- | --- | --- | --- | --- | --- |
| Overall | 0.617 ± 0.305 | 0.550 ± 0.248 | –0.067 | (–0.081,<br>–0.052) | <0.001 |
| T2DM | 0.617 ± 0.305 | 0.545 ± 0.239 | –0.071 | (–0.089,<br>–0.053) | <0.001 |
| Non-T2DM | 0.617 ± 0.305 | 0.558 ± 0.261 | –0.058 | (–0.084,<br>–0.033) | <0.001 |

**Note:** Values are mean ± SD. PFQI = protein (%E) ÷ fat (%E). STARCH 2014 statistics are from Joshi et al. (2014). STARCH 2014 reported only an overall PFQI, so the T2DM and non-T2DM comparisons use the same reference value. Comparisons are descriptive between independent cross-sectional cohorts. **Abbreviations:** PFQI, Protein–Fat Quality Index; SD, standard deviation; T2DM, type 2 diabetes mellitus.

## 4. Discussion

### 4.1 Principal Findings

The present multicentre cross-sectional study provides contemporary evidence on macronutrient intake and carbohydrate quality among Indian adults with and without T2DM. Consistent with previous national observations, carbohydrates remained the predominant source of dietary energy, whereas protein contributed a relatively modest proportion of total energy intake, and dietary fibre intake remained suboptimal. Simple carbohydrates accounted for approximately one-fifth of total carbohydrate intake, indicating scope for improvement in carbohydrate quality. Participants with T2DM exhibited only modest differences in macronutrient distribution and carbohydrate quality compared with those without T2DM, while dietary patterns were broadly similar across the four geographic regions, despite statistically significant but small regional differences.

### 4.2. Comparison with Previous Literature

The present findings are consistent with the STARCH 2014 study, which identified carbohydrates as the predominant contributor to total energy intake among Indian adults with and without T2DM (Joshi et al., 2014). The I-STARCH 2025 cohort showed a modestly lower carbohydrate contribution and a higher fat contribution than STARCH 2014, yet carbohydrates remained the principal energy source, indicating that this dietary pattern persists nationally. As STARCH 2014 and I-STARCH 2025 are independent cross-sectional cohorts from different periods, this comparison should be read descriptively rather than as evidence of temporal change. These findings are also concordant with the ICMR–INDIAB studies and the ICMR–NIN Dietary Guidelines for Indians, which describe Indian diets as cereal-carbohydrate dominant with comparatively low fibre and protein density (Anjana et al., 2023, 2025; ICMR–NIN, 2024). Similar carbohydrate-dominant patterns have been reported in other Indian multicentre studies, suggesting this pattern is widespread across regions and population groups rather than confined to specific geographic or clinical subgroups (Green et al., 2016; Hemalatha et al., 2023; Mohan et al., 2009).

The present findings also align with international evidence describing the ongoing nutrition transition, characterized by increasing consumption of refined carbohydrates, processed foods, and energy-dense diets (Ma et al., 2024; Malik & Hu, 2022). Systematic reviews and prospective cohort studies have consistently shown that cardiometabolic risk is influenced not only by the quantity of carbohydrate consumed but also by its quality, particularly the balance between refined carbohydrates, whole grains, and dietary fiber (Reynolds et al., 2019; Ma et al., 2024; Forouhi et al., 2018; Jannasch et al., 2017). In this context, the greater proportion of simple carbohydrates observed in the I-STARCH 2025 cohort relative to the published STARCH 2014 cohort reinforces the importance of improving carbohydrate quality, alongside maintaining an appropriate macronutrient balance, as a priority for nutritional guidance in Indian adults. Because food-source analyses were not performed, a higher simple carbohydrate share may reflect multiple dietary sources, including sweetened beverages, desserts, packaged snacks, and refined carbohydrate-containing foods, rather than any single dietary component (Malik & Hu, 2022).

### 4.3. Mechanistic Interpretation

The observed dietary patterns are consistent with established mechanisms linking carbohydrate quality to metabolic health. Diets rich in refined carbohydrates may contribute to greater post-prandial glycaemic excursions, hyperinsulinaemia, and insulin resistance (Reynolds et al., 2019; Forouhi et al., 2018; Lambadiari et al., 2020). Inadequate dietary fibre has also been associated with impaired glucose homeostasis through reduced short-chain fatty acid production and altered satiety signalling (Chambers et al., 2018; Niero et al., 2023). In the present cohort, protein contributed a relatively modest proportion of total energy intake, while dietary fat contributed a greater proportion than in the historical STARCH cohort, indicating a shift in macronutrient composition rather than a clear improvement in overall dietary quality (Mohan et al., 2023a; Gulati & Misra, 2017). Collectively, these mechanisms reinforce the importance of improving carbohydrate quality and dietary fibre intake, while maintaining adequate protein intake, rather than focusing solely on carbohydrate quantity, for cardiometabolic health in Indian adults.

### 4.4. Interpretation of Differences Between Participants With and Without T2DM

Participants with T2DM exhibited only modest differences in macronutrient distribution and carbohydrate quality compared with those without T2DM. Several factors may account for these observations in the present study. Individuals with established diabetes are more likely to have received dietary counselling and intentionally modify their dietary habits following diagnosis, which may contribute to lower carbohydrate intake and greater emphasis on healthier carbohydrate choices (American Diabetes Association Professional Practice Committee, 2025; Maghsoudi & Azadbakht, 2012). Consistent with this interpretation, a greater proportion of participants with T2DM in the present study reported intentionally restricting their diet in the preceding year than those without T2DM. Differences in age may also have influenced habitual dietary intake, while medication use, disease duration, and comorbidities, which were not comprehensively captured in this survey, represent potential sources of unmeasured confounding. Furthermore, self-reported dietary assessments are susceptible to reporting bias, particularly among individuals with diagnosed diabetes who may be more aware of recommended dietary practices (Livingstone & Black, 2003; Poslusna et al., 2009). Given the cross-sectional design, these findings should not be interpreted as evidence that diabetes status causes the observed dietary differences.

### 4.5. Geographic Variation in Dietary Patterns

Dietary patterns were broadly similar across the participating geographic zones, with carbohydrates remaining the predominant source of dietary energy and only modest variations observed in macronutrient distribution and carbohydrate quality across North, South, East, and West India. However, because participants were recruited from specialty clinics rather than through population-based sampling, these findings should not be interpreted as evidence of regional uniformity at the population level. Dietary practices across India remain highly heterogeneous, reflecting differences in staple foods, cultural traditions, and socioeconomic factors (Green et al., 2016; ICMR–NIN, 2024). Therefore, the similarities observed in the present study should be interpreted within the context of the participating centers.

### 4.6. Public Health Implications

The present findings suggest that dietary guidance for Indian adults should emphasize carbohydrate quality rather than simply reducing carbohydrate quantity. Given the modest fibre intake and high simple-carbohydrate contribution observed, priorities include greater consumption of whole grains, legumes, millets, and minimally processed foods, alongside reduced free and simple sugar intake, consistent with current national recommendations (ICMR– NIN, 2024; Saboo et al., 2022; RSSDI 2025 Consensus Group, 2025). Protein contributed a relatively modest proportion of total energy intake in this cohort, reinforcing the importance of promoting adequate intake from high-quality protein sources within balanced diets (Saboo et al., 2022). Because dietary patterns were broadly similar across diabetes status and geographic zones, evidence-based dietary counselling in both clinical and community settings may support sustained improvements beyond carbohydrate restriction alone. Behavioral strategies informed by neuromarketing principles may further support healthier population-level food choices (Anthuvan et al., 2024). Future public health policy should prioritize carbohydrate quality, fibre density, adequate protein intake, and minimally processed foods, although these approaches were not directly evaluated in the present study.

### 4.7. Clinical Implications for Weight Management

The present findings carry additional clinical relevance given the increasing use of anti-obesity pharmacotherapy. Protein intake in this cohort (12.8%E) was below the 15–20%E range recommended by RSSDI–ICMR-NIN guidelines (RSSDI 2025 Consensus Group, 2025). National dietary data further indicate that cereals contribute most protein intake in Indian diets, suggesting both protein quantity and quality may be suboptimal (Athinarayanan et al., 2026; Hemalatha et al., 2023). This is particularly relevant for individuals undergoing intentional weight loss, including those on anti-obesity pharmacotherapy, who may need closer attention to protein intake to preserve lean body mass during weight reduction. Current evidence supports combining adequate protein intake with resistance exercise to optimize body composition and functional outcomes during weight loss (American Diabetes Association Professional Practice Committee, 2025; RSSDI 2025 Consensus Group, 2025; Madhu et al., 2025; Neeland et al., 2024; Verma et al., 2022). Culturally relevant approaches, such as structured bite-count reduction, have also supported sustainable weight loss in Indian adults (Anthuvan & Salis, 2025). Although beyond this study’s scope, these considerations reinforce the importance of integrating protein adequacy with contemporary obesity management.

### 4.8. Strengths

This study had several strengths. It draws on a large, contemporary sample of 1,104 Indian adults with and without T2DM recruited from 29 healthcare centers across 14 states, providing broad geographic coverage spanning the north, south, east, and west of the country. Dietary intake was assessed using three non-consecutive 24-hour dietary recalls administered by trained investigators, and nutrient values were derived using harmonized coding based on the ICMR-NIN Food Composition Tables, which supports methodological consistency across participating centers. The inclusion of adults with and without T2DM in the same multicenter study enabled a direct comparison of macronutrient intake and carbohydrate quality using a common methodology. While geographic diversity strengthens the dataset relative to previous single-region studies, the clinic-based convenience sample should not be interpreted as nationally representative.

### 4.9. Limitations

This study has several limitations. The cross-sectional design precludes causal inference, and the convenience-based recruitment of participants from specialty clinics limits the generalizability of the findings to the wider Indian population. Participant enrolment varied across centers, and dietary intake was based on self-reported 24-hour recalls, making the data susceptible to recall bias, social desirability bias, and potential under-reporting, while seasonal variation in dietary intake could not be fully captured. Residual confounding from unmeasured factors, including medication use, disease duration, socioeconomic status, and lifestyle behaviors, may have influenced the observed findings. Although T2DM status was determined using medical records, medication history, self-report where necessary, and biochemical data where available, biochemical confirmation was not uniformly available for all participants, and misclassification of diabetes subtypes cannot be completely excluded. Nutrient estimation may also have been affected by matching some branded foods to generic food-composition entries and by regional food substitutions within the ICMR-NIN Food Composition Tables when exact matches were unavailable, potentially introducing minor measurement errors. Comparisons with the published STARCH 2014 cohort should be interpreted as contextual historical comparisons because the studies differed in sampling framework, participant characteristics, and dietary assessment procedures, and because harmonized individual-level historical data were unavailable, requiring comparisons based on published summary statistics rather than participant-level data. Finally, the Diet-Quality Index (DQI) and Protein–Fat Quality Index (PFQI) used in this study are investigator-derived exploratory indices that have not undergone external validation.

### 4.10. Future Research

Future research should build on these findings through population-based sampling and longitudinal dietary surveillance to determine whether the patterns observed in this clinic-based cohort reflect broader national dietary trends and how they evolve over time. The Diet Quality Index (DQI) and Protein–Fat Quality Index (PFQI) require further validation, including external validation against objective biomarkers such as HbA1c, lipid profile, body mass index, waist circumference, and hepatic markers, to establish their clinical utility and predictive value for cardiometabolic outcomes. Future studies should also incorporate food-source analyses, distinguish free sugars from intrinsic sugars to better characterize carbohydrate quality, and employ seasonally repeated dietary recalls to capture temporal variations in dietary intake. Such work will strengthen dietary assessment methods and improve the applicability of these indices for clinical practice, nutrition surveillance, and future dietary policies.

## 5. Conclusions

This large multicenter study of Indian adults with and without T2DM found that carbohydrates remained the predominant source of dietary energy, while dietary fiber intake remained low, protein contributed a relatively modest proportion of total energy intake, and simple carbohydrates accounted for a substantial proportion of total carbohydrate intake. Differences in macronutrient distribution and carbohydrate quality between participants with and without T2DM and across the participating geographic regions were generally modest. Compared descriptively with the published STARCH 2014 cohort, the findings suggest a less favorable carbohydrate quality profile, characterized by a lower contribution of carbohydrates to total energy intake, a higher contribution from fat, and a greater proportion of simple carbohydrates. Because these comparisons were based on independent cross-sectional cohorts, they should be interpreted cautiously. Collectively, these findings support shifting nutritional guidance from reducing carbohydrate quantity alone to improving carbohydrate quality by increasing dietary fiber and minimally processed carbohydrate sources while limiting free and simple sugars. Future population-based and longitudinal studies are needed to confirm these findings and to inform evidence-based dietary strategies for cardiometabolic health in India.

## Supplementary Materials

The following supporting information can be downloaded at: [MDPI website link], Table S1: STROBE-Nut Checklist for the I-STARCH-1 Study; File S1: Dietary Survey Questionnaire; Figure S1: Macronutrient distribution and carbohydrate quality according to diabetes status; Figure S2: Geographic distribution of macronutrient intake across the I-STARCH 2025 cohort.

## Author Contributions

Conceptualization, S.K. and T.A.; methodology, S.K., N.K., N.D. and S.S.S.; investigation, S.S.S. and S.G.; data curation, T.A. and S.S.S.; formal analysis, T.A.; writing—original draft preparation, T.A. and S.S.S.; writing—review and editing, S.K., N.K., N.D. and S.G.; supervision, S.K.; project administration, T.A., S.S.S. and S.G.; funding acquisition, S.G. All authors have read and agreed to the published version of the manuscript. S.K. accepts full responsibility for the integrity of the work as a whole, had access to the aggregated study data, and takes responsibility for the decision to submit the manuscript for publication.

## Institutional Review Board Statement

The study was conducted in accordance with the Declaration of Helsinki, the Indian Council of Medical Research (ICMR) National Ethical Guidelines for Biomedical and Health Research Involving Human Participants, and the applicable regulatory requirements. The study was approved by the Institutional Ethics Committee of the Shah Lifeline Hospital & Heart Institute (Approval No. SLH-IEC/2025/006, approved on January 17, 2025).

## Informed Consent Statement

Written informed consent was obtained from all participants before enrolment.

## Data Availability Statement

The data presented in this study are available on request from the corresponding author. The data are not publicly available because they contain information that could compromise participant privacy, and because participant consent did not provide for open data deposition. The completed STROBE-Nut checklist and the structured dietary survey questionnaire are provided in the Supplementary Materials. Participating-centre enrolment details are available from the corresponding author on request. Requests are subject to ethics approval, institutional permission, and execution of an appropriate data-use agreement.

## Data Availability

De-identified data supporting the findings of this study are available from the corresponding author upon reasonable request for non-commercial academic use, subject to institutional approvals and a data-use agreement.

## Acknowledgments

The authors gratefully acknowledge the site investigators and their research teams across the 29 participating centers for their contributions to participant recruitment, study coordination and data collection: Dr Ajish T. P. (Kollam), Dr Anamika Samant (Mumbai), Dr Anwar Jamal (Kolkata), Dr Arun Karthik (Coimbatore), Dr D. K. Srivastava (Patna), Dr G. Shanmugasundar (Chennai), Dr Hamid Ashraf (Aligarh), Dr Hiranmoy Paul (Siliguri), Dr Jagdish Mulchandani (New Delhi), Dr K. Venugopal Reddy (Vijayawada), Dr Kalpesh Kavar (Rajkot), Dr Manash Baruah (Guwahati), Dr Neelakshi Deka (Guwahati), Dr Neeta Deshpande (Belagavi), Dr Nilesh Detroja (Rajkot), Dr Nitin Kapoor (Coimbatore), Dr P. K. Gupta (Chennai), Dr P. Sasikanth Reddy (Hyderabad), Dr R. K. Singh (Samastipur), Dr Raka Sheohore (Raipur), Dr Rupam Choudhary (Guwahati), Dr S. Paramesh (Bengaluru), Dr Saqib Ahmad Khan (Lucknow), Dr Saurabh Arora (Ludhiana), Dr Shaibal Guha (Patna), Dr V. K. Dhandania (Ranchi), Dr Yogesh Varge (Akola), Dr Yogesh Yadav (Dehradun) and Dt. Sheryl S. Salis (Mumbai). The authors thank the Nurture Health Solutions team for the investigator training and standardization of dietary recall procedures. They acknowledge the Department of Computer Science, Sardar Patel University, Anand, Gujarat, for technical guidance during the development of the computational workflow used for dietary data harmonization. The authors also thank USV Private Limited and Mediclin Clinical Research Organisation for their operational support. An AI-assisted computational tool was used to support food code harmonization and nutrient computation; the tool did not generate text, interpretations, or participant-level conclusions, and all outputs were reviewed and verified by trained investigators before analysis. The custom code used for dietary data harmonization and analysis is available from the corresponding author upon reasonable request and will be deposited in an appropriate public repository upon manuscript acceptance.

Patients and the public were not involved in the design, conduct, reporting, or dissemination of this study. This manuscript was prepared in accordance with the STROBE-Nut reporting guidelines, and the completed STROBE-Nut checklist is available from the corresponding author upon reasonable request. All exploratory analyses, including the Diet Quality Index and Protein– Fat Quality Index, were explicitly identified as exploratory analyses.

